# Coronavirus protective immunity is short-lasting

**DOI:** 10.1101/2020.05.11.20086439

**Authors:** Arthur W. D. Edridge, Joanna Kaczorowska, Alexis C. R. Hoste, Margreet Bakker, Michelle Klein, Maarten F. Jebbink, Amy Matser, Cormac M. Kinsella, Paloma Rueda, Maria Prins, Patricia Sastre, Martin Deijs, Lia van der Hoek

## Abstract

In the current COVID-19 pandemic a key unsolved question is the duration of acquired immunity in recovered individuals. The recent emergence of SARS-CoV-2 precludes a direct study on this virus, but the four seasonal human coronaviruses may reveal common characteristics applicable to all human coronaviruses. We monitored healthy subjects over a time span of 35 years (1985-2020), providing a total of 2473 follow up person-months, and determined a) the time to reinfection by the same seasonal coronavirus and b) the dynamics of coronavirus antibody depletion post-infection. An alarmingly short duration of protective immunity to coronaviruses was found. Reinfections occurred frequently at 12 months post-infection and there was for each virus a substantial reduction in antibody levels as soon as 6 months post-infection.

## MAIN TEXT

SARS-CoV-2 is a novel coronavirus responsible for an ongoing pandemic. Its rapid transmission is most probably caused by the fact that the virus entered a grossly naive, thus highly susceptible, human population, combined with the capacity of the virus to transmit during the asymptomatic phase of infection. Since no pharmaceutical interventions are universally available, current policies to limit the spread of SARS-CoV-2 revolve around containment, social distancing, and the assumption that recovered patients develop protective immunity. The duration of protection will impact not only the overall course of the current pandemic, but also the post-pandemic period. To date, no concrete evidence of reinfection by SARS-CoV-2 is available, nor is any example of reinfection by SARS-CoV-1 or MERS-CoV, yet this is likely influenced by the recent emergence of SARS-CoV-2 and the limited scale of the SARS-CoV-1 and MERS-CoV epidemics. It is generally assumed that reinfection by coronaviruses can occur, however, this is based on only one study with experimental infection in volunteers using a cultured coronavirus (HCoV-229E) as inoculum with a 12 month interval^1^. Susceptibility to natural reinfection by coronaviruses has thus not been investigated. If they occur, reinfections will probably be dictated by two variables: exposure to the virus, and the duration of sustained immunity^2^.

It is not possible to investigate SARS-CoV-2 reinfections currently, since we are in an early phase of the pandemic, yet the seasonal coronaviruses may serve as a model. There are four species of seasonal coronaviruses, HCoV-NL63, HCoV-229E, HCoV-OC43, and HCoV-HKU1. All are associated with mostly mild respiratory tract infections, however, the four viruses are genetically and biologically dissimilar. Two belong to the genus *Alphacoronavirus*, and two to the genus *Betacoronavirus*. The viruses use different receptor molecules to enter a target cell, and based on receptor distribution they do not all enter the same epithelial cell type in the lungs^3^. Given this variability, the seasonal coronaviruses are the most representative virus group from which to conclude general coronavirus characteristics, particularly common denominators like dynamics of immunity and susceptibility to reinfection.

The aim of this study is to investigate the duration of coronavirus protection from reinfections. Examining reinfections by testing for the virus in respiratory material requires respiratory sample collection from volunteers during each common cold symptom and, because reinfections can be asymptomatic, also during symptom-free periods, for years in a row. These type of sample collections are difficult to obtain. The alternative is measuring rises in antibodies to a virus (serology), as an indicator of recent infection. This option is applicable, as it has been determined that IgG levels to seasonal coronavirus 1) only increase after successful infection, 2) also rise with asymptomatic infection, and 3) do not increase after unsuccessful viral challenge^1^. Blood samples collected at regular intervals in cohort studies following volunteers for decades can subsequently deliver the essential material to investigate serology-based reinfection dynamics. In theory, virus-neutralization tests may seem the best serology assay when protective immunity is investigated, however, there are serious limitations. First, there is no cell line facilitating replication of HCoV-HKU1, and therefore neutralization tests cannot be done for this virus. Second, the only available HCoV-229E and HCoV-OC43 cultured virus strains are from the 1960’s and lab-adapted, which may not be proper representatives of wild type viruses. Instead, measuring antibody levels directed to viral proteins using for instance ELISA, will allow reinfection-testing for all 4 seasonal coronaviruses. In that case, a careful choice for the antigen has to be made, considering the trade-off between sensitivity and cross-reactivity in a serological assay. Although the Spike protein elicits neutralizing antibodies, it is the least conserved within a seasonal coronavirus species^4^, and decreasing the sensitivity to detect infections. In contrast, the N protein, and specifically its C terminal region (NCt), is significantly more conserved and has been found as the most immunogenic, specific and sensitive protein to monitor seasonal coronavirus infections^5–9^.

### Infection dynamics

From a prospective cohort study following adult males (see M&M and ^10^), ten subjects, who participated since the start of the study and at least 10 years of follow up, were selected. These participants did not report any serious illness that could have influenced their immunity (see description of the cohort in the M&M). Follow-up of subjects with blood collection and storage started in 1985 and, besides a gap in follow up between 1997 and 2003, continued for most subjects until 2020 at regular intervals (every 3 months prior to 1989, and every 6 months afterwards). The cumulative period at which subjects were continuously followed (i.e. < 400 days interval between consecutive samples) totaled more than 200 person-years: 2473 months. At start of the study, subject age ranged from 27 to 40 years; by the end of follow-up, subjects were 49 to 66 years old.

Coronavirus infections were determined by measuring fold changes in optical density (OD) in NCt antibody recognition between two subsequent visits (shown for one individual in **Fig. 1A** and for all subjects in **supplementary Fig. S1**). We measured first the natural fluctuation among consecutive visits in measles virus antibodies for all 10 subjects, assuming measles did not occur during follow up, as all subjects were vaccinated during childhood. Fold changes in antibody OD for measles virus ranged between 0.85 and 1.28 (**Fig. 1B**). A threshold for coronavirus-infection was next determined using the distribution of the OD fold change, assuming that during most intervals no coronavirus infection occurred and infections therefore appear as outliers. **Figure 1B** shows that ≥1.4 fold OD rises were outliers (**Fig. 1B**). We next determined whether these serological infection criteria could be confirmed with self-reported influenza like illnesses (ILI) in the interval directly preceding the rise in antibodies. Indeed, reporting of ILI-symptoms coincided with a ≥1.4 rise in antibodies (Fisher’s exact test p=0.031, **Supplementary table S1**). Finally we compared ELISA results of HCoV-NL63 with neutralization titers for HCoV-NL63 for two subjects (#5 and #7, three infections). The infections showed an increase in neutralization titer accompanying the ≥1,4 fold rise in NCt antibodies (**Fig. 1C**).

**Figure 1.**
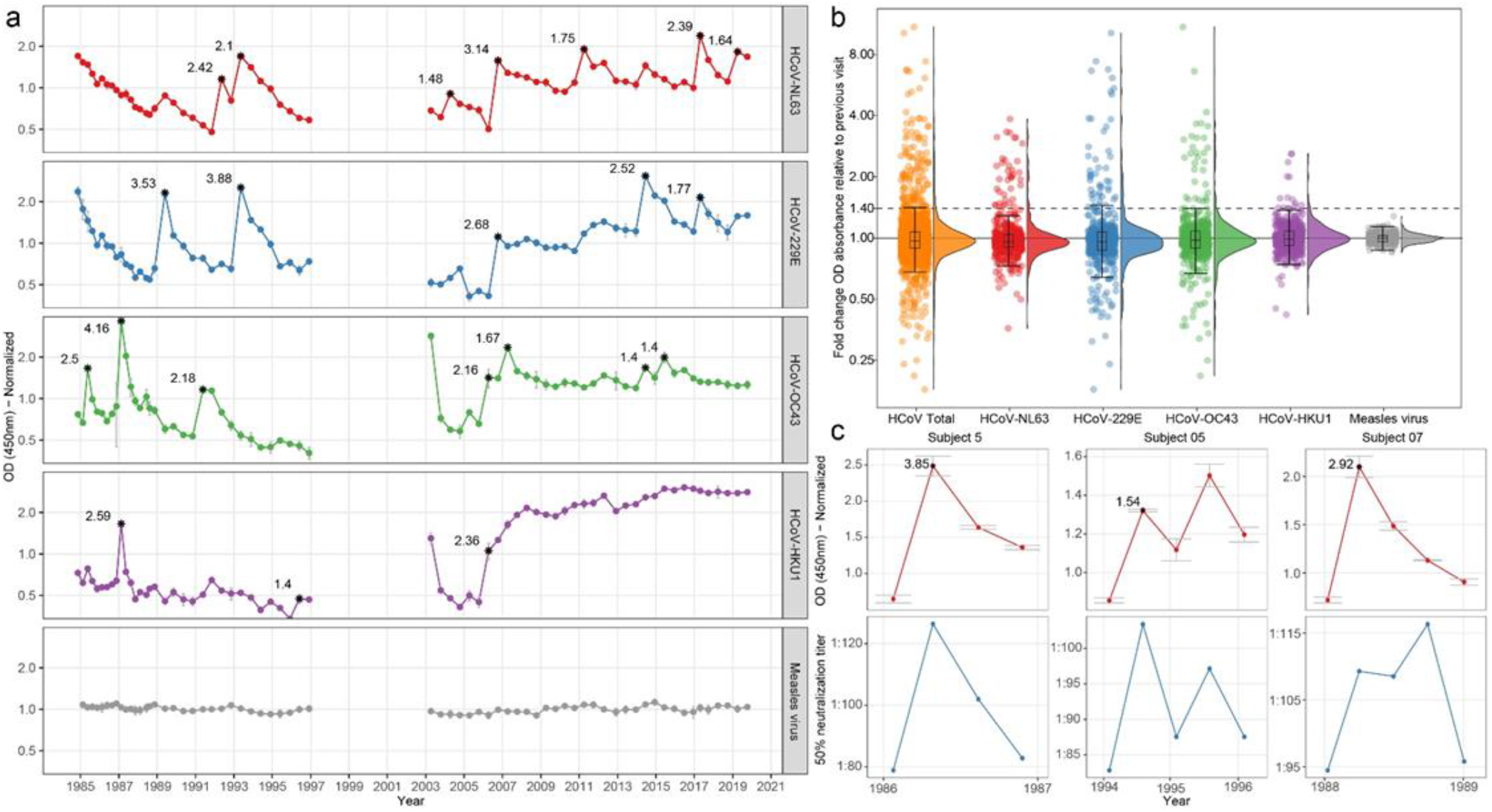
Antibody dynamics for seasonal coronaviruses and measles virus in time. **(A)** Antibody dynamics of subject 9. Connected dots indicate follow up intervals <400 days. Asterisks signify visits classified as infections, the number adjacent to the asterisk describes the observed fold change in optical density of the ELISA. **(B)** Distribution of OD fold changes between sequential visits. Boxplots show median values with the interquartile range (IQR), the whiskers extend up to the range after which samples are considered outliers (1.5 x IQR below the first or above the third quantile). In red: HCoV-NL63; blue: HCoV-229E; green: HCoV-OC43; purple: HCoV-HKU1; orange: total seasonal coronaviruses; grey: measles virus. (**C)** Antibody reactivity to HCoV-NL63-NCt associates with neutralization of HCoV-NL63. In red: antibody dynamics measured by ELISA; blue: neutralization titers.

A total of 132 events, ranging from 3 to 22 per subject, were classified as coronavirus infections (**Table 1**). Median reinfection times of 33 (IQR 18 – 60), 31 (IQR 15 – 42), 27 (IQR 21 – 49), and 46 (IQR 36 – 68) months were found for HCoV-NL63, HCoV-229E, HCoV-OC43 and HCoV-HKU1, respectively, and 30 (IQR 18 – 54) months for all viruses combined (**Fig. 2A**). There was no statistically significant difference between the infection interval lengths of the individual viruses (Kruskal-Wallis test, P=0.74). In a few cases, re-infections occurred as early as 6 months (two times for HCoV-229E and one time for HCoV-OC43) and 9 months (two times for HCoV-NL63). The most frequent observed reinfection time was 12 months. For reinfections occurring as early as 6 months, we observed no reduction in antibodies between infections (**Fig. 2A**, white circles), yet reinfections times > 6 months showed reductions in antibody levels between two infections (visible as peaks in **Fig. 1A and Supplementary Fig. S1**).

**Figure 2.**
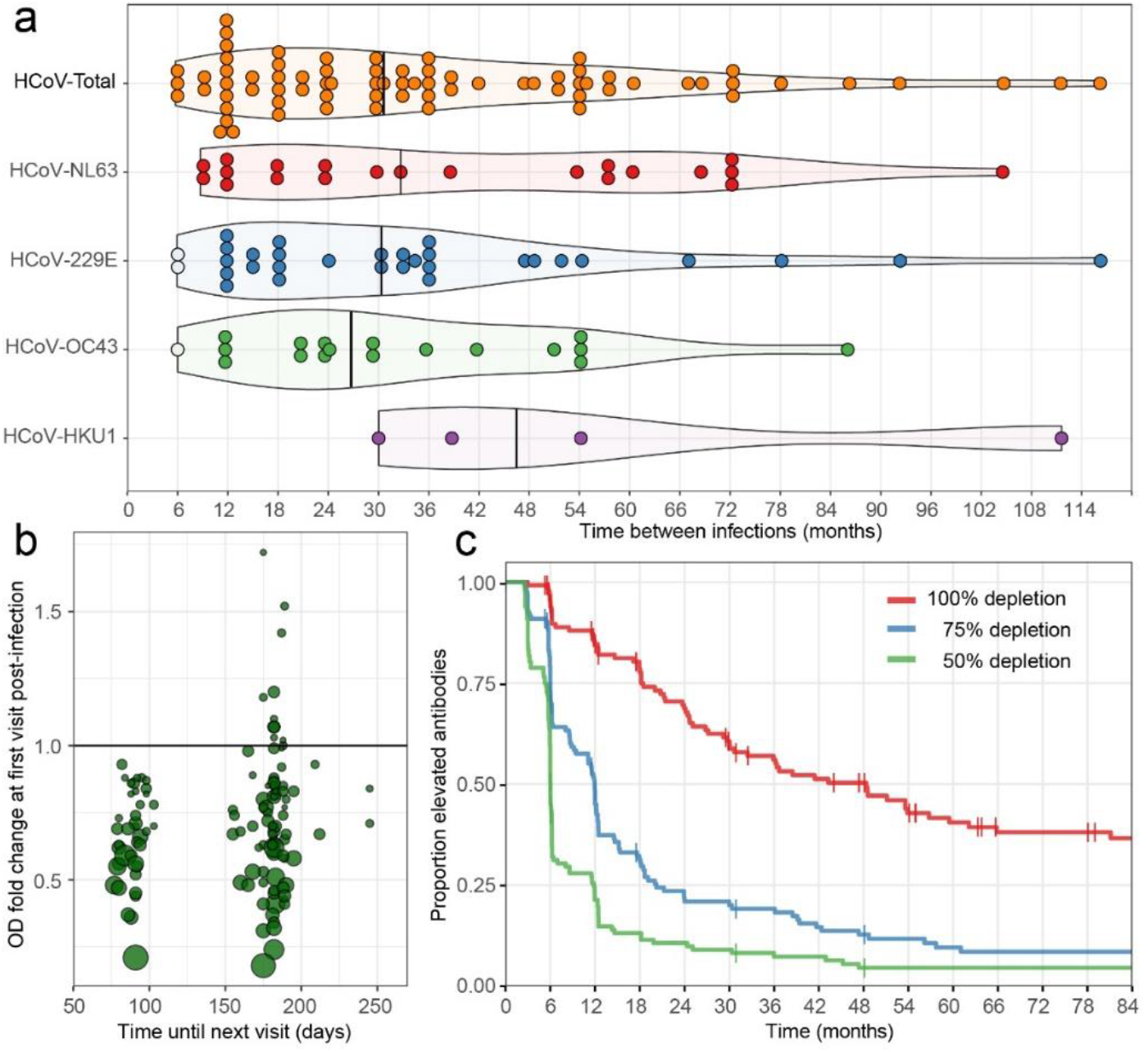
Infection and reinfection characteristics, and dynamics of waning antibodies for seasonal coronaviruses. **(A)** The interval time between reinfections. White dots indicate reinfections for which no intermediate decrease in antibody levels could be observed. Black vertical lines describe median reinfection times. (**B**) Changes in antibody levels post-infection relative to the follow-up interval duration. Each circle represents an infection. The x-axis describes the time until the next follow-up visit post-infection. The y-axis describes the change in antibody level at the subsequent visit. Larger circles represents a higher ratio-rise in antibody levels at the initial infection. The horizontal line indicates the border between increases (>1.0) or decreases (<1.0) in antibody levels at the next study visit. (**C**) Kaplan-Meier curve showing decline of antibodies post infection (100%, 75% and 50%), details in M&M.

**Table 1.** Study subjects and seasonal coronavirus infections during follow up.

| Subject | Year |  | Age |  | Continuous follow-up period |  | Coronavirus Infections |  |  |  |  |
| --- | --- | --- | --- | --- | --- | --- | --- | --- | --- | --- | --- |
|  | Start | End | Start | End | Months | Years | Total* | NL63 | 229E | HKU1 | OC43 |
| 1 | 1985 | 2017 | 32 | 64 | 265 | 22.1 | 14 | 4 | 3 | 1 | 6 |
| 2 | 1985 | 2019 | 30 | 64 | 310 | 25.9 | 15 | 5 | 6 | 3 | 1 |
| 3 | 1985 | 2020 | 29 | 64 | 340 | 28.3 | 6 | 2 | 3 | 0 | 1 |
| 4 | 1985 | 2010 | 33 | 59 | 230 | 19.2 | 22 | 3 | 12 | 2 | 5 |
| 5 | 1985 | 2010 | 27 | 53 | 232 | 19.3 | 9 | 5 | 3 | 1 | 0 |
| 6 | 1985 | 1997 | 37 | 49 | 144 | 12.0 | 3 | 1 | 1 | 0 | 1 |
| 7 | 1985 | 2003 | 32 | 49 | 138 | 11.5 | 13 | 3 | 5 | 1 | 4 |
| 8 | 1986 | 2014 | 34 | 62 | 256 | 21.3 | 10 | 3 | 5 | 0 | 2 |
| 9 | 1985 | 2010 | 40 | 75 | 342 | 28.6 | 22 | 7 | 5 | 3 | 7 |
| 10 | 1985 | 2011 | 35 | 60 | 233 | 19.4 | 18 | 4 | 6 | 2 | 6 |
| Total |  |  |  |  | 2473 | 205.6 | 132 | 37 | 49 | 13 | 33 |

The ability to detect short-term reinfections is limited by the sampling interval. Importantly though, no reinfection was observed at the first subsequent follow-up visit after a 3 month interval (**Fig. 2A**). We did observe several reinfections at subsequent visits with a 6-month interval, suggesting that reinfections within 6 months do not occur. To more closely examine this we also looked at the <1.4 fold changes in antibodies directly after an infection, under the assumption that antibody levels which do not decrease adequately post-infection may be a sign of another infection, comparing 3 month interval data or 6 month interval data. As shown in **Fig. 2B**, only fold changes below 1 were found after infection with every 3 month-sampling, and we can therefore safely conclude that the earliest confident time point for reinfection is 6 months.

### Antibody dynamics after infection

Protective immunity may last shorter than the measured time until reinfection because a reinfection also requires exposure to a virus. Therefore, depletion of antibody levels may be a better marker of waning immunity. Although antibodies to the N-protein by itself are not neutralizing, they can be regarded as a representative of the total of antibodies (see comparisons with neutralization **Fig. 1C**). We analyzed the dynamics of the decline of NCt-antibodies post-infection, by calculating the time until a 50%, 75%, or full return of antibody levels to baseline (pre-infection antibody levels) occurred. The majority of patients lost 50% of their NCt-protein-antibodies after 6 months, 75% after a year, and completely returned to baseline 4 years post-infection (**Fig. 2C and Supplementary Fig. S1**).

### Simultaneous infections

Although our ELISA tests using the C-terminal part of the N protein were cautiously designed to be specific for each individual virus, we cannot rule out that a certain degree of antibody cross-reactivity occurred. We therefore investigated how often infections coincided, since cross-reactivity may have led to false labeling of infections. We observed that simultaneous infections with an alphacoronavirus (HCoV-NL63 or HCoV-229E) alongside betacoronaviruses (HCoV-HKU1 or HCoV-OC43) were rare, however, we did see that infections by the betacoronaviruses HCoV-OC43 and HCoV-HKU1 often coincided (38.5%, **Table 2**). Likewise, for the alphacoronaviruses, HCoV-229E infections coincided with HCoV-NL63 infections in 59.5% of the cases, and *vice versa* in 44.9% of the cases. Hence, there is a risk that we overestimated the number of infections and thus reinfections when antibody rises occurred for two coronaviruses of the same species. We therefore re-analyzed the data with a more stringent definition of infection, including only the strongest antibody rise induced by a *Betacoronavirus* or *Alphacoronavirus* at a given time point. Under this definition we still found infection intervals comparable to the original data (**supplementary Fig. S2**), with minimum infection intervals as short as 6 months and frequent reinfections at 12 months, although the number of reinfections was obviously reduced.

**Table 2.** Coinciding coronavirus infections

| Infection | Simultaneous infections (%) |  |  |  |
| --- | --- | --- | --- | --- |
|  | HCoV-NL63 | HCoV-229E | HCoV-OC43 | HCoV-HKU1 |
| HCoV-NL63 | NA | 44.9 | 3.0 | 7.7 |
| HCoV-229E | 59.5 | NA | 18.2 | 23.1 |
| HCoV-OC43 | 2.7 | 12.2 | NA | 38.5 |
| HCoV-HKU1 | 2.7 | 6.1 | 15.2 | NA |

### Broadly recognizing coronavirus antibodies

In theory, antibodies induced by coronavirus infections may have broad coronavirus-recognizing characteristics. To examine this, we performed an additional ELISA on all 10 subjects, this time using the complete N protein of SARS-CoV-2, to allow detection of broadly recognizing antibodies. To exclude that the detection of these kind of antibodies were false positives, e.g. because they were directed to the his-tag on our coronavirus N-proteins, we also performed a control ELISA with a his-tagged HIV-1 envelope protein. One subject (#2) showed non-specific recognition, since the his-tagged HIV-1-SOSIP protein was also recognized (**Supplementary Fig. S3**). Two other subjects showed broadly recognizing antibodies, most likely induced by infections with *Alphacoronavirus* and *Betacoronavirus* during the same interval (subjects 9 and 10, **Supplementary Fig. S3**). Of note, it does not seem that broadly recognizing antibodies are broadly protective as HCoV-NL63, HCoV-229E, and HCoV- OC43 infections occur in the presence of the broadly recognizing antibodies (**Supplementary figure S1**).

### Coronavirus infections in changing seasons

To date it is uncertain whether SARS-CoV-2 will share the same winter prevalence peak that is observed for seasonal coronaviruses in non-equatorial countries. However, it is important to consider that winter preference of seasonal coronaviruses has only been determined by testing respiratory samples of people that experienced disease^11^. Sampling and storage is therefore dictated by having symptoms and not by study protocol. If coronavirus spread continues unabated in summer, yet people rarely display symptoms and are therefore not sampled, infections will remain undetected. Our serological study is unique because it avoids this sampling bias. The Netherlands has a typical temperate climate, and our study samples were collected at regular intervals. The sampling of each subject were randomly distributed throughout the year, and, because of the 3 or 6 months regimen of visits, samples were collected throughout all seasons. Consequently, we can for the first time visualize the seasonality of coronavirus infections in an unbiased manner. We estimated the prevalence of infection onset for each month for all infections detected in this study (see Supplementary methods for analysis details). As shown in **Fig. 4**, the spring and summer months May, June, July, August and September show the lowest prevalence of infections for all four seasonal coronaviruses (Wilcoxon signed-rank test, p=0.005).

**Fig. 4.**
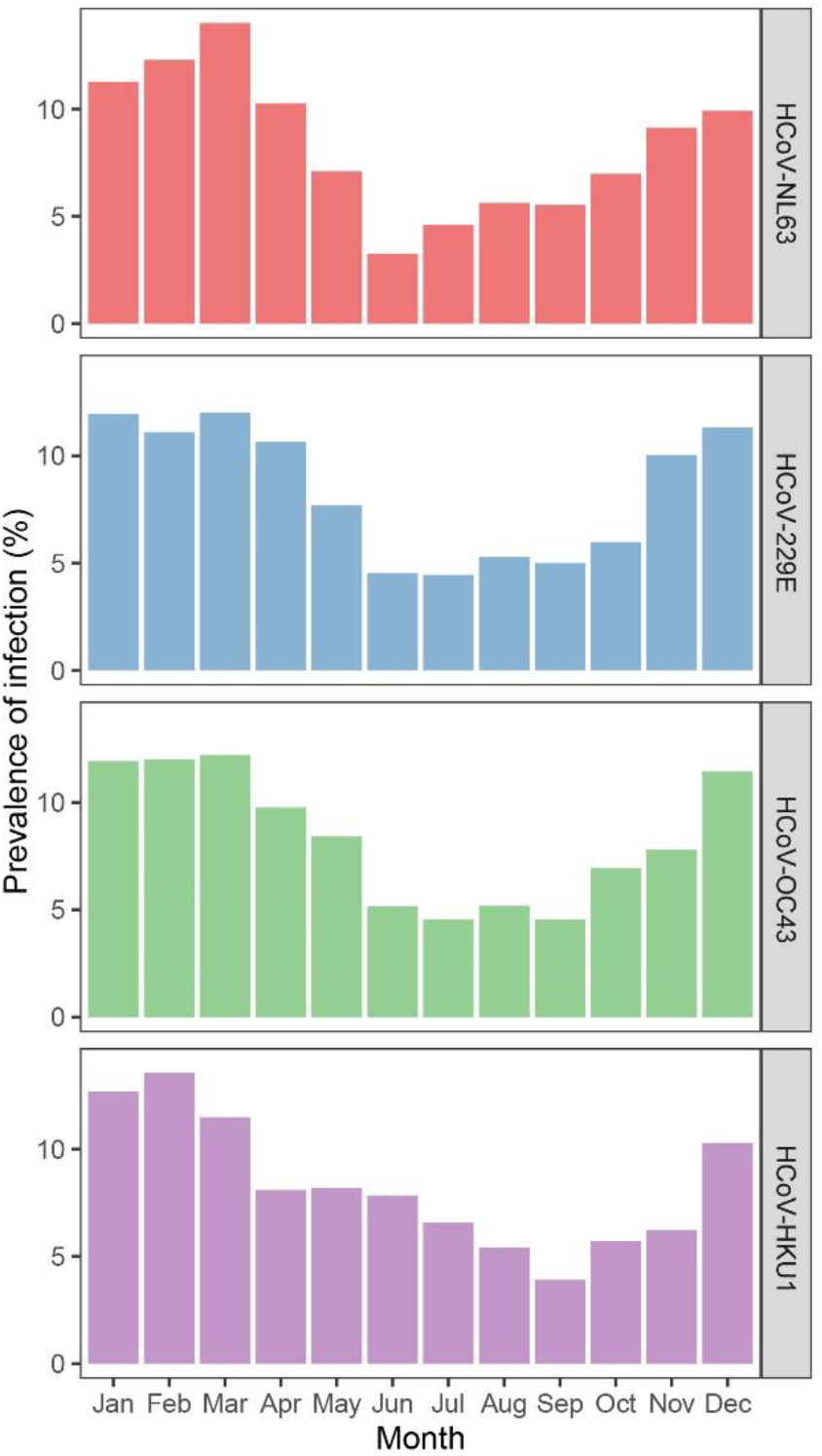
Seasonality of infections. The prevalence of infection of the four seasonal coronaviruses across different months, the prevalence per month is shown as a percentage of the total number of infections per coronavirus.

## DISCUSSION

We show, for the first time, that reinfections with all seasonal coronaviruses occur by natural infection. The majority of reinfections occurred within 3 years. However, this time span between infections does not indicate that an individual’s protective immunity lasts for the same period of time, as reinfection is also dependent on re-exposure. In fact, based on the minimum infection intervals and the dynamics of antibody waning that we observed, protective immunity may last as little as 6 to 12 months. Recently Kissler *et al*. modeled the protective immunity and reinfection dynamics HCoV-OC43 and HCoV-HKU1 and estimated a 45 week period of protective immunity^12^. Our serological study confirms this prediction.

When we view our findings in light of the current control actions taken for SARS-CoV-2, it is clear that coronavirus reinfection risk is key to public health policy. Here we reveal a risk that in the near future, serology based tests that measure previous infections for SARS-CoV-2 using the N-protein may have limited use if that infection has occurred >1 year prior to sampling. Our study also shows that herd immunity may be challenging due to rapid loss of protective immunity. It was recently suggested that recovered individuals should receive a so-called “immunity passport”^13^ which would allow them to relax social distancing measures and provide governments with data on herd immunity levels in the population. However, as protective immunity may be lost by 6 months post infection, the prospect of reaching functional herd immunity by natural infection seems very unlikely.

We noticed three subjects to carry antibodies recognizing SARS-CoV-2 N protein at certain time points. It is unlikely that they had been infected with a SARS-CoV-2-like virus in 1985 (subject # 10), 1992 (subject #2), or 2006 (subject #9), and we therefore suggest that broadly recognizing antibodies may have been induced by coinciding infections of an *Alpha-* and a *Betacoronavirus* (in our subjects HCoV-HKU1 and HCoV-NL63). To explore this finding we looked at the genetic distance and consequently amino acid differences in the structural protein of the various coronaviruses (**supplementary Table S2**). SARS-CoV-2 N protein has only 32% and 34% identity on the amino acid level with the N protein of HCoV-OC43 and HCoV-HKU1 respectively, and only 26% and 24% identity with HCoV-NL63 and HCoV-229E respectively. Similarly, the distance between *Alphacoronavirus* and *Betacoronavirus* N protein is large (only 24% to 26% amino acid identity). Still, we cannot exclude the presence of conserved (conformational) epitopes in HCoV-HKU1 and HCoV-NL63 N protein that may result in a more broadly acting antibody response, due to simultaneous exposure in concurrent infections. Additional screening, including more subjects, is required for confirmation.

We were not able to sequence the virus genome during infection. In theory, strain variation could play a role in susceptibility to reinfection. HCoV-NL63, HCoV-OC43, and HCoV-HKU1 all show different co-circulating genetic clusters^4,14,15^. The situation is even more complicated for HCoV-229E. Sylvia Reed has shown that experimental re-infection of volunteers is not successful when the same strain of HCoV-229E is used, yet successful when heterologous strains are used^16^. Strangely, no major genetic subtypes are known for HCoV-229E^17,18^. As HCoV-NL63 was not known in the 1980’s, and culture characteristics differed between the HCoV-229E strains of Reed, the so-called heterologous strains may actually have been HCoV-NL63^16,19^. A study on protective immunity would therefore ideally allow sequencing of re-infecting strains from respiratory material; however, this is intractable in a natural infection study because virus shedding in reinfections can be as short as one day, and respiratory sampling schemes would be extremely cumbersome for volunteers^1^. Another limitation of the study is that the subjects in our study were all males. For COVID-19, and also HCoV-NL63, men have a higher incidence of disease^20^, and it is therefore of interest to determine the dynamics of protective immunity also in a cohort of healthy women.

In conclusion, seasonal human coronaviruses have little in common, apart from causing common cold. Still, they all seem to induce a short-lasting immunity with rapid loss of antibodies. This may well be a general denominator for human coronaviruses.

## Data Availability

All data are available upon request from the authors

## Acknowledgments

The authors gratefully acknowledge the Amsterdam Cohort Studies (ACS) on HIV infection and AIDS, a collaboration between the Public Health Service of Amsterdam, the Amsterdam UMC of the University of Amsterdam, Sanquin Blood Supply Foundation, Medical Center Jan van Goyen, and the HIV Focus Center of the DC-Clinics. It is part of the Netherlands HIV Monitoring Foundation and financially supported by the Center for Infectious Disease Control of the Netherlands National Institute for Public Health and the Environment. The authors thank all ACS participants for their contribution, as well as the ACS study nurses, data-managers, and lab technicians. This work was supported by a grant from the European Union’s Horizon 2020 research and innovation programme, under the Marie Skłodowska-Curie Actions grant agreement no. 721367 (HONOURs), Amsterdam UMC funding connected to HONOURs, and the Amsterdam UMC PhD scholarship of A.W.D. Edridge.

## Author contributions

A.W.D.E: Conceptualization, Writing – original draft, review and editing, Investigation, Visualization, Formal Analysis, Validation; J.K.: Investigation, Writing – original draft, review and editing, A.C.R.H.: Resources; M.B.: Investigation, Resources; M.K.: Investigation; M.F.J.: Investigation, Methodology; A.M.: Formal Analysis; C.M.K.: Writing – original draft, review and editing, Formal Analysis; P.R.: Resources; M.P.: Resources; P.S.: Resources; M. D.: Investigation, Methodology; L.v.d.H.: Conceptualization, Writing – original draft, Writing - review and editing, Supervision.

## Competing interests

Authors declare no competing interests.

## Data and materials availability

All data is available in the main text or the supplementary materials. Materials are available for study via the open and ongoing Amsterdam Cohort Studies (ACS) on HIV among MSM: https://www.eed.amsterdam.nl/beleid-onderzoek/projecten/amsterdamse-cohort/

## Notes

### Competing Interest Statement

The authors have declared no competing interest.

### Author Declarations

This study is approved by the Medical Ethics Committee of the Amsterdam University Medical Center of the University of Amsterdam, the Netherlands (MEC 07/182). Participation is voluntary and without incentive. Written informed consent of each participant was obtained at enrollment.

